# Vitamin B12 Absorption in a Community is ‘Continuously’ Distributed and Influences Response to Long-term Oral Supplementation

**DOI:** 10.1101/2025.07.18.25331760

**Authors:** Chittaranjan S Yajnik, Elaine C Rush, Dattatray S Bhat, Rucha SH Wagh, Omkar A Deshmukh, Rasika Ladkat, Lindsay D Plank, Souvik Bandopadhyay, Caroline H Fall, Helga Refsum, Urmila S Deshmukh

## Abstract

**Background:** Insufficient dietary intake and/or reduced gastrointestinal absorption lead to low vitamin B_12_ (B-12) status. In a secondary data analysis of the results of the B-12 absorption (CobaSorb) test and B-12 supplementation trial in a B-12 insufficient Indian rural community, we explored characteristics of B-12 absorption and its association with post-supplementation plasma B-12 concentration.

**Objective:** To study i) the distribution of plasma holo-transcobalamin (holoTC) response during the CobaSorb test, and ii) determinants of plasma B-12 response to long-term, oral supplementation with ‘physiological dose’ B-12.

**Methods:** The participants (parents and children in the Pune Maternal Nutrition Study) first underwent a B-12 absorption study Subsequently, they participated in a 12-month-long, double-blind RCT of daily oral B-12 (0, 2, or 10 µg) and folic acid (0 or 200 µg). Circulating B-12 was measured at baseline and after 4 and 12 months of supplementation. A linear mixed-effect model was used to study the predictors of B-12 absorption and response to supplementation.

**Results:** Three hundred and thirteen participants included children (n=109, 57 boys, mean age 9y, weight 21.9 kg, BMI 13.6 kg/m^2^), mothers (n=108, 30y, 47.7 kg, 19.3 kg/m^2^), and fathers (n=96, 37y, 59.3 kg, 21.4 kg/m^2^). The plasma holoTC response during the absorption test was continuously distributed and was negatively associated with weight and positively with the dose of B-12. Response to long-term B-12 supplementation was positively predicted by absorption test response, dose of B-12, length of supplementation, and compliance.

**Conclusions:** The continuous distribution of plasma holoTC response during the absorption test supports a graded absorption of B-12. CobaSorb protocol could be used to unravel the intricacies of B-12 absorption.

**Clinical Trial Registry number and website where it was obtained:** ISRCTN59289820. DOI: https://doi.org/10.1186/ISRCTN59289820 https://www.isrctn.com/

## Introduction

Vitamin B_12_, a water-soluble vitamin, cannot be synthesized in humans and is available through diet and possibly from the intestinal microbiota. Dietary vitamin B_12_ (B-12) is mostly sourced from animal products. Worldwide, B-12 insufficiency and deficiency are increasingly driven by reduced intake of animal products, particularly red meat, and in aging populations, additionally by food-B12 (cobalamin) malabsorption [1, 2]. The interpretation of biomarkers of B-12 status requires an understanding of the process of B-12 absorption and metabolism.

Orally consumed B-12 attaches with salivary carrier proteins, is transferred to the intrinsic factor (IF), which mediates its intestinal absorption. It is transported to tissues, attached to two proteins, and is an essential component of many metabolic pathways. Up to 70% of absorbed vitamin is transported by haptocorrin, which is stored only by the liver; the remaining 30% is transported by transcobalamin II, which is delivered to peripheral tissues and is available for cellular metabolism [1, 3]. The latter is called holo-transcobalamin (holoTC) or ‘active B-12’ [4]. Tissue metabolism includes cytoplasmic methylation reactions, which generate methionine from homocysteine and provide S-adenosyl methionine (SAM), a universal methyl donor. Vitamin B12 is also a coenzyme for a mutase enzyme, which converts methylmalonic acid (MMA) to propionic acid, which then enters the mitochondria for energy production. Elevated circulating total homocysteine (tHcy) and MMA concentrations indicate reduced tissue availability of B-12. Elevated tHcy may also reflect deficiencies of other ‘methyl’ vitamins (folate, pyridoxine, riboflavin) while elevated MMA is usually considered a specific biomarker of B-12 deficiency [1–4].

Conventional ideas about B-12 absorption were based on the Schilling test, which was mostly performed with patients suspected of suffering from pernicious anaemia and resulted in binary reports of ‘absorber’/ ‘non-absorber’ [5]. Schilling’s test used a radioactive isotope of cobalt and is not available now. Stable isotope labelled vitamin is being used to investigate B-12 absorption in specialized research laboratories [6]. The CobaSorb test investigates the absorption of B-12 from the gut and its bioavailability by measuring the rise in circulating levels of holoTC after oral administration of ‘physiological’ doses of B-12. It may be used in routine clinical practice. There are only a few reports of this measure with healthy controls and little information on the distribution of absorption characteristics in B12-deficient populations [7].

India has a high prevalence of low plasma B-12 status, attributed to the low intake of animal-origin foods. In our cross-sectional and longitudinal cohort studies, we have found a high prevalence of low B-12 status (<150 pmol/L coupled with hyperhomocysteinemia in 21 to 70 %) throughout the lifecourse [8–11].

We used the CobaSorb protocol of Bor *et al* [12] to investigate B-12 absorption in 313 participants of the Pune Maternal Nutrition Study (parents and children) by measuring the rise in circulating holoTC concentration using physiological doses of oral B-12 (10 and 2 µg x 3 doses) [13]. This study [13] showed that the 24-hour rise in holoTC after an oral challenge with B-12 was normal in >90% of participants based on Bor’s criteria. The same parents and children subsequently participated in a randomized controlled trial of physiological doses of B-12 and folic acid (daily 2 or 10 µg B-12, with and without 200 µg folic acid), for 12 months [14]. Circulating B-12 concentrations were measured at baseline, and 4 and 12 months after starting the supplementation.

These datasets [13, 14] provided an opportunity to undertake a secondary analysis to study the distribution of the increase in holoTC in the CobaSorb test and to examine the relationships between the increase in holoTC during the absorption test and the response of circulating B-12 concentrations 4 and 12 months after supplementation.

## Methods

### Study Setting and Participants

Briefly, the Pune Maternal Nutrition Study (PMNS) is a community-based, prospective birth cohort established during 1993-94 in six villages near Pune [15]. The original PMNS enrolled 814 pregnant women for the study of maternal nutrition and fetal growth. An additional 153 women from the same population were enrolled (extended PMNS) after the completion of the original study to standardize ultrasonic measurements of early fetal growth in this undernourished population. These women had similar characteristics to the original cohort.

Parents and children in the original and the extended PMNS have been followed up regularly to study physical growth and the evolution of type 2 diabetes and cardiovascular disease. One of the important nutritional findings in the PMNS was the high rates of low B-12 concentrations and hyperhomocysteinemia in this largely vegetarian population [11]. We performed a B-12 absorption study (CobaSorb test: rise in circulating concentrations of holoTC in response to 3 physiological oral doses of B-12, 6 h apart) to rule out absorption defects in this population [13]. This was followed by a pilot trial of physiological dose B-12 and folic acid supplementation to improve one-carbon metabolism [14], in anticipation of our intergenerational trial [16] to reduce the primordial risk of diabesity in the population. The KEM Hospital Ethics Committee approved both studies (KEMHRC/VSP/Dir Off/EC/065; Project No. 067).

This is a secondary analysis of the data generated in the absorption study [13] and the pilot B-12-folic acid supplementation trial [14] in the participants of the extended PMNS cohort. The details of the B-12 absorption protocol [13] and B-12 supplementation trial [14] are summarized in **Figure 1**. This secondary analysis investigates the characteristics of ‘absorption performance’ (rise in circulating holoTC concentrations in response to oral B-12 during the CobaSorb test) to test the hypothesis that the B-12 absorption performance is continuously distributed rather than binary (‘yes/no’). We further investigated whether the B-12 absorption performance was associated with the response of circulating B-12 concentrations during long-term supplementation.

**Figure 1:**
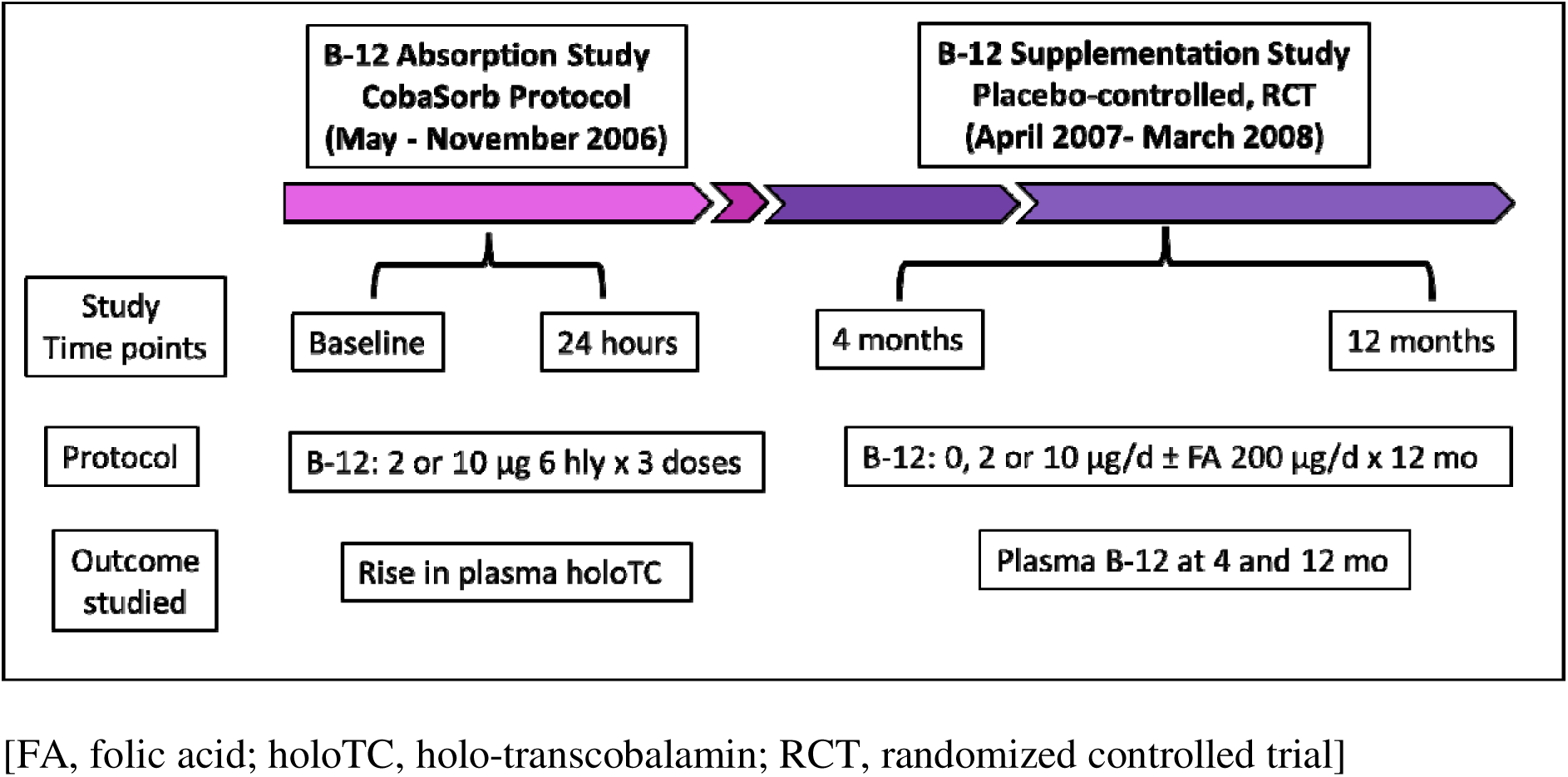
Schematic presentation of B-12 Absorption and Supplementation studies.

### Absorption study protocol

One hundred and nineteen families, from the follow-up in the extended PMNS, were invited to participate (May to November 2006); 109 families participated [13]. The participants were admitted overnight for the absorption study in the Diabetes Unit, KEM Hospital & Research Centre, Pune. Standardised food and facilities were provided. Demographic details and anthropometric measurements were recorded. A fasting blood sample was collected (baseline), and a 10 µg (64 families) or 2 µg (44 families) B-12 (cyanocobalamin) capsules were administered under supervision every 6 h x 3 times. A fasting blood sample was collected the following morning (post-dose sample) to measure the rise in circulating holoTC concentration.

### Supplementation study

The same participants were enrolled in this randomized, stratified, placebo-controlled longitudinal trial with a factorial design (April 2007 – March 2008; ISRCTN59289820, Protocol No. 079877/Z/06/Z) [14]. The participants were randomized to receive one of three daily doses of B-12 (0, 2, or 10 µg daily) and two doses of folic acid (0 or 200 µg), forming six groups (B_0_F_0,_ B_2_F_0_, B_10_F_0_, B_0_F_200_, B_2_F_200,_ and B_10_F_200_). Randomization was computer-based. The unit of randomization was the family, making it a cluster-randomized trial. Stratification was by the children’s baseline plasma B12 concentrations (below and above the median value of 188 pmol/L). The duration of supplementation was 12 months, and a non-fasting blood sample was collected at 4 and 12 months after starting supplementation to measure circulating B-12 concentrations. Compliance with B-12 supplements was assessed monthly by counting the unused capsules and as a percentage of the capsules consumed.

### Anthropometry and Laboratory Measurements

Height was measured to the nearest 0.1cm using a wall-mounted stadiometer (CMS Instruments, London, UK), and body weight to the nearest 0.05 kg (Conveigh, Electronic Instruments Ltd, Mumbai, India). Plasma B-12 and folate were measured by microbiological assays using a colistin sulfate-resistant strain of *Lactobacillus leishmannii* [17, 18] and a chloramphenicol-resistant strain of *Lactobacillus casei I* [19, 20], with inter-batch CVs of <8% and <7%, respectively. Plasma holoTC was measured using magnetic beads (microspheres) with an immobilized monoclonal antibody specific for human transcobalamin, followed by the conventional microbiological assay developed for cobalamin estimation [21, 22].

The combined B-12 marker (cB12) was estimated, using plasma B-12, total homocysteine, and folate concentrations, using Fedosov’s method [23]. B-12 adequacy status at baseline, 4, and 12 months was estimated using the cut points of cB12 defined for epidemiological purposes, viz. B-12 adequacy: cB12 >-0.5, transitional B-12 status: cB12 ≥-2.5 and ≤-0.5, low B-12 status: cB12 <-2.5 [23].

### Statistical methods

Data are presented as mean ± SD, or as median and 25^th^–75^th^ percentile when not normally distributed. The Shapiro-Wilk test was used to test normality. Variables not normally distributed were transformed to the natural logarithm before analysis (weight, plasma B-12, folate, and holoTC). Correlation and linear mixed effects regression analyses were used to explore associations and predictors.

#### B-12 Absorption Study

We studied the distribution of the rise in holoTC concentration during the absorption study by plotting it graphically. We investigated the contribution of the following predictors to the rise in plasma holoTC: body weight, sex, and dose of B-12 (2 or 10 µg), using linear mixed-effects regression analysis.

#### B-12 Supplementation trial

Predictors of circulating B-12 concentrations at 4 and 12 months included: absorption performance (the change in holoTC during the absorption test, adjusted for the dose of B-12), the dose of B-12 (0, 2, or 10 µg), and folic acid (0 or 200 µg) in the supplementation trial, and the compliance and length of supplementation. We used linear mixed-effects regression analyses for this purpose. All statistical analyses were carried out using R software (R ver. 4.4).

## Results

### Recruitment and participant flow

The consort diagram (**Figure 2**) shows the flow of participants in the two studies and exclusions or drop-outs. A total of 313 individuals participated in the B-12 absorption study; 311 had all measurements. Three hundred participants were randomized in the supplementation trial; 291 had all measurements at 4 months and 287 at 12 months.

**Figure 2:**
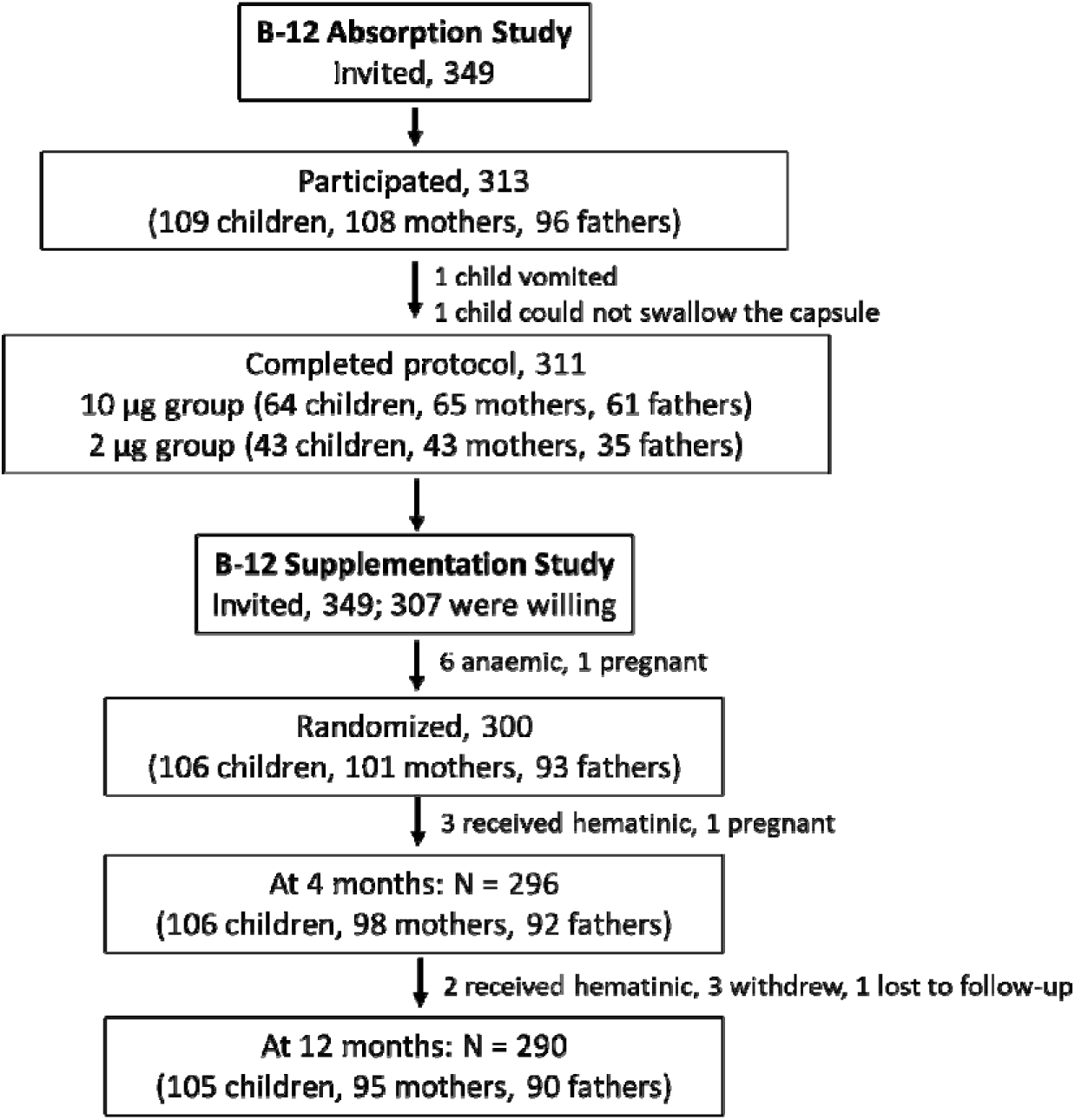
Participant flow in the B-12 Absorption and Supplementation studies.

### Baseline characteristics

Children (n=109; 57 boys, 52 girls) were 9 years old, mothers (n=108) were 30 years old, and fathers (n=96) were 37 years old. Children weighed 21.9 kg, mothers 47.7 kg, and fathers 59.3 kg. Low plasma B-12 (<150 pmol/L) concentrations were present in 27% of children, 48% of mothers, and 72% of fathers.

### Absorption Study

#### Plasma B-12 and holoTC concentrations

As already reported (13), there was a satisfactory rise in holoTC concentrations in the majority of participants. The median rise in holoTC concentrations was 42.0 pmol/l from baseline for the 10 µg dose (55.0 in children, 39.0 in mothers, 39.0 in fathers), and 23.0 pmol/l for the 2 µg dose (39.0 in children, 19.0 in mothers, 10.0 in fathers). Overall, plasma holoTC increased 4.8-fold with the 10 µg dose and 2.2-fold with the 2 µg dose; the rise was greater in children than in the parents (**Table 1** and **Figure 3**).

**Figure 3:**
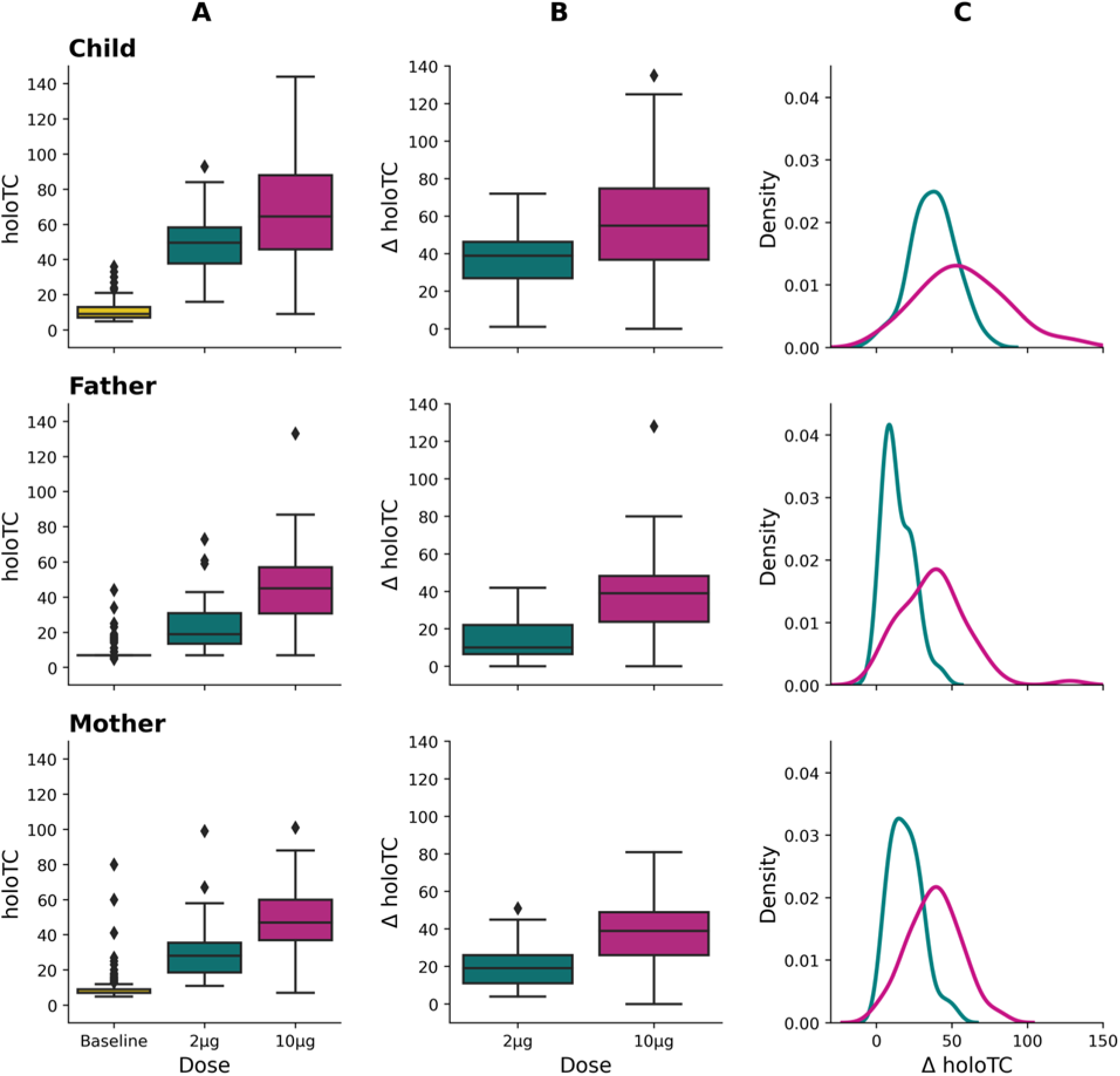
Results of the CobaSorb Absorption Study in children, fathers, and mothers after 3 doses of 2 and 10 µg B-12. A: Box plots of baseline and 24 hr plasma holoTC. B: Rise in plasma holoTC from the baseline. C: Density plots of the rise in plasma holoTC.

**Figure 4:**
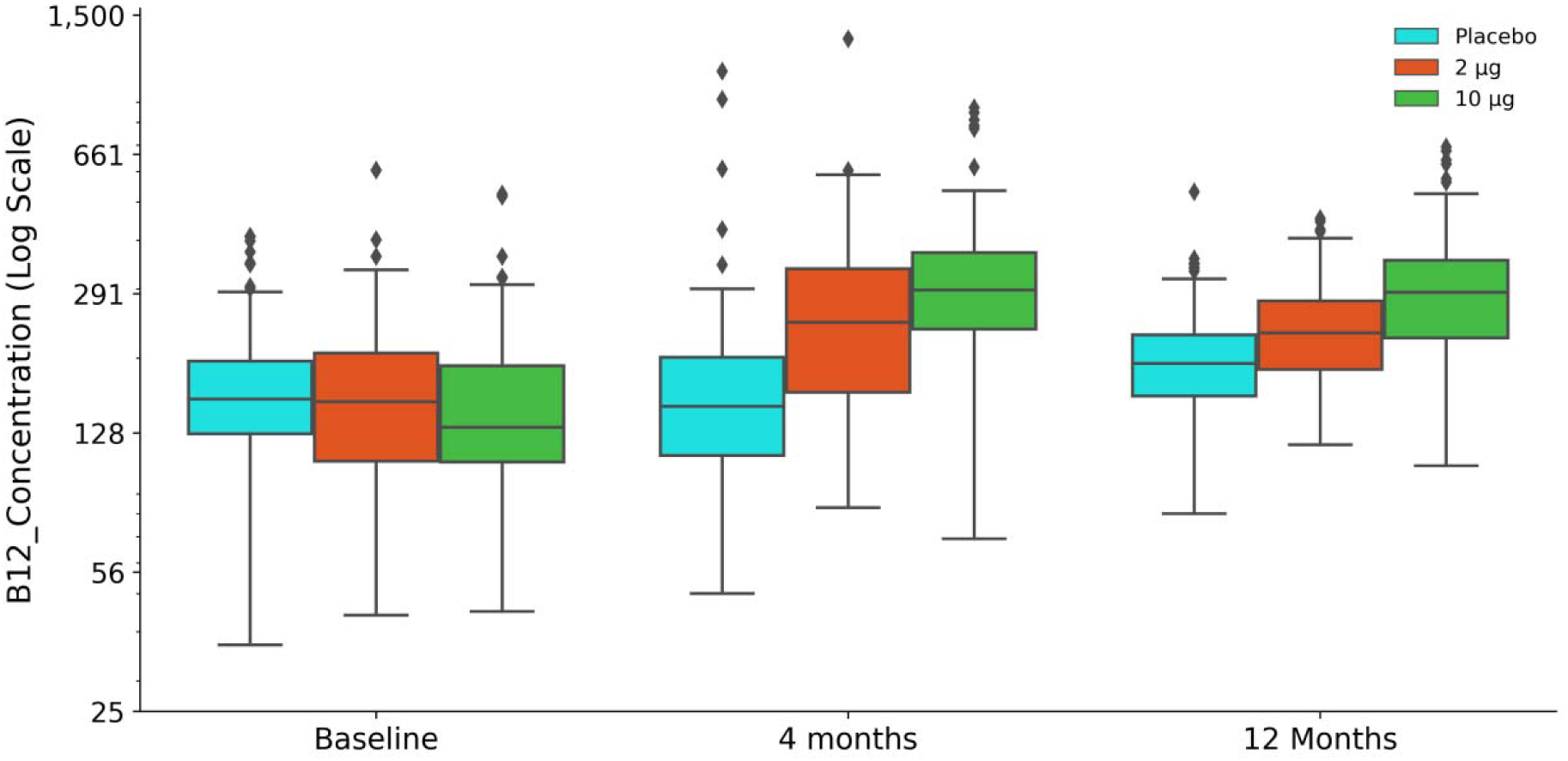
Supplementation Study: Box plots of plasma B-12 at baseline, 4, and 12 months in the three B-12 supplemented groups (0, 2, and 10 µg B-12).

**Table 1.** Absorption Study: Response of plasma holo-TC to the B-12 Absorption protocol in 2 and 10 µg B-12 groups.

|  | 10 µg group |  |  | 2 µg group |  |  |
| --- | --- | --- | --- | --- | --- | --- |
|  | Children | Father | Mother | Children | Father | Mother |
| *N* | 64 | 60 | 65 | 43 | 35 | 43 |
| Baseline plasma | 9.0 | 7.0 | 7.0 | 7.5 | 7.0 | 7.0 |
| holo-TC, *pmol/L* | (7.0, 10.8) | (5.0, 7.0) | (7.0, 10.0) | (7.0, 15.8) | (7.0, 9.0) | (7.0, 9.0) |
| 24-hr plasma | 64.5 | 45.0 | 47.0 | 49.5 | 19.0 | 28.0 |
| holo-TC, *pmol/L* | (45.3, 88.0) | (30.3, 57.0) | (36.5, 60.5) | (37.3, 58.8) | (13.0, 32.0) | (18.0, 36.0) |
| Rise of plasma | 55.0 | 39.0 | 39.0 | 39.0 | 10.0 | 19.0 |
| holo-TC from | (36.3, 76.3) | (23.3, 48.8) | (25.5, 50.5) | (27.0, 46.8) | (6.0, 22.0) | (11.0, 26.0) |
| baseline, *pmol/L* |  |  |  |  |  |  |

Numbers are median (25<sup>th</sup>, 75<sup>th</sup> percentile).

Using Bor et al.’s 2004 criterion (rise in plasma holoTC <15% and <15 pmol/L after 3 doses of 9 µg B-12) [12], 10% of participants (10 fathers, 5 mothers, and 4 children) in the 10 µg group were classified as poor absorbers. In a subsequent study by Bor et al., including patients with inherited B-12 malabsorption (Imerslund-Grasbeck syndrome or intrinsic factor deficiency), a rise of ≥ 6 pmol/L of holoTC was proposed as a cut point to define ‘adequate’ B-12 absorption (100% sensitivity and 72% specificity) [24]. Using this cut point, 3.7% of participants in our study (3 fathers, 2 mothers, and 2 children) could be classified as inadequate absorbers.

Figure 3 shows the results of the CobaSorb Absorption Study for children, fathers, and mothers. Panel A presents a box plot of baseline and 24-hour holoTC concentrations in the 2 and 10 μg groups, while panel B illustrates a box plot of the rise in holoTC concentrations for the same dosage groups. Panel C displays density plots of the increase in plasma holoTC at 24 hours following three doses of 2 and 10 µg. The distributions are nearly bell-shaped, with a slight right skew due to a few outliers (Shapiro-Wilk test, p<0.001 for plasma holoTC at baseline, 24 hours, and the change). These distributions were normalized using a log transformation for further analysis. The density plots can be interpreted in terms of the peak (average), width (variability), and shifts (comparing the effects of 2 and 10 µg B-12 doses). The peaks shift further to the right in children compared to parents, indicating a higher average rise for both doses. The width is smaller for the 2-µg dose than for the 10-µg dose, suggesting less variability, and the curves for the 10-µg dose are shifted to the right compared to the 2-µg dose, demonstrating a greater rise. The predominant characteristic of the density plots shows that the distribution is continuous and not binary or bimodal.

In multiple regression analysis, the increase in holoTC concentration exhibited an inverse association with body weight and a direct association with the doses of B-12 (10 or 2 µg) (**Table 2**). The most significant effect was observed for the B-12 dose.

**Table 2:** Absorption Study: Predictors of the rise in plasma holoTC concentration.

|  | Beta (S.E.) | *P* |
| --- | --- | --- |
| Weight (kg) | -0.565 (0.060) | <0.0001 |
| Sex: Female | -2.416 (2.070) | 0.244 |
| Baseline plasma holoTC (pmol/l) | 0.098 (0.147) | 0.507 |
| B12 dose: 10µg (reference: 2µg) | 19.881 (2.680) | <0.0001 |
| R<sup>2</sup> | 0.464 |  |

### Supplementation Study

Plasma B-12 concentrations increased clinically significantly in those who received B-12 supplements, and plasma folate concentrations increased in those who received folic acid (**Table 3**). At 12 months, the median B-12 rise was 1.9-fold (95% CI: 1.8–2.2) from baseline in the 10 µg group, compared to 1.5-fold (95% CI: 1.3–1.7) in the 2 µg group. Using the combined B-12 marker (cB12), the B-12 adequacy status remained stable in the placebo group (baseline 46.0 % and 12 months 50.0 %), while it improved from 41.5 to 73.3 % in the 2 µg group, and from 35.7 to 85.7 % in the 10 µg group (p < 0.001 for the B-12 groups) (**Table 4**).

**Table 3.** Supplementation Study: Plasma B12, folate, and homocysteine responses in the six groups after 4 and 12 months of intervention.

|  | B<sub>0</sub>F<sub>0</sub><br>n=48 | B<sub>2</sub>F<sub>0</sub><br>n=46 | B<sub>10</sub>F<sub>0</sub><br>n=47 | B<sub>0</sub>F<sub>200</sub><br>n=52 | B<sub>2</sub>F<sub>200</sub><br>n=47 | B<sub>10</sub>F<sub>200</sub><br>n=51 |
| --- | --- | --- | --- | --- | --- | --- |
| Plasma B<sub>12</sub> (pmol/L) |  |  |  |  |  |  |
| Baseline | 154<br>(127, 196) | 150<br>(96, 205) | 133<br>(100, 189) | 159<br>(132, 200) | 158<br>(109, 207) | 130<br>(110, 201) |
| 4 mo | 155<br>(113, 195) | 235<br>(148, 331) *** | 298<br>(210, 455) *** | 149<br>(112, 214) | 259<br>(173, 340) *** | 298<br>(245, 357) *** |
| 12 mo | 192<br>(159, 225) ** | 216<br>(184, 281) *** | 293<br>(215, 364) *** | 198<br>(160, 247) ** | 242<br>(196, 286) *** | 297<br>(224, 338) *** |
| Plasma folate (nmol/L) |  |  |  |  |  |  |
| Baseline | 16.4<br>(12.5, 23.4) | 15.6<br>(12.3, 20.2) | 16.8<br>(13.3, 21.5) | 15.7<br>(12.8, 20.1) | 15.8<br>(12.6, 17.9) | 17.2<br>(13.3, 22.9) |
| 4 mo | 15.5<br>(11.0, 19.5) | 14.5<br>(10.7, 18.1) * | 16.0<br>(11.4, 18.6) * | 28.0<br>(21.1, 47.9) *** | 31.4<br>(20.8, 45.7) *** | 28.2<br>(21.4, 40.2) *** |
| 12 mo | 15.4<br>(10.2, 21.1) | 13.3<br>(10.2, 16.2) *** | 12.9<br>(10.5, 16.3) *** | 28.2<br>(17.1, 38.8) *** | 24.5<br>(17.1, 33.5) *** | 24.8<br>(16.5, 31.1) *** |
| Plasma total homocysteine (μmol/L) |  |  |  |  |  |  |
| Baseline | 11.5<br>(8.8, 22.1) | 14.5<br>(10.0, 21.3) | 14.4<br>(10.2, 23.6) | 11.5<br>(8.8, 17.7) | 12.9<br>(9.6, 18.0) | 13.2<br>(9.4, 18.8) |
| 4 mo | 13.6<br>(9.9, 23.6) | 11.5<br>(8.0, 18.3) * | 10.1<br>(8.3, 16.3) *** | 11.4<br>(8.4, 17.0) | 9.6<br>(7.3, 12.2) *** | 9.2<br>(7.3, 14.0) *** |
| 12 mo | 13.8<br>(10.5, 23.0) | 11.9<br>(8.8, 18.0) ** | 9.2<br>(7.5, 14.1) *** | 13.6<br>(9.2, 16.8) | 9.6<br>(8.0, 12.6) *** | 9.3<br>(7.6, 12.4) *** |
| Compliance at 12 mo (%) | 82 | 88 | 89 | 81 | 83 | 85 |

Median (25<sup>th</sup>, 75<sup>th</sup> percentiles) unless specified. \* difference from baseline concentration within groups by linear mixed models. \**P* < 0.05, \*\**P* < 0.01, \*\*\**P* < 0.001. [The groups are: B<sub>0</sub>F<sub>0</sub> = placebo, B<sub>2</sub>F<sub>0</sub> = B-12 2 μg and folic acid 0, B<sub>10</sub>F<sub>0</sub> = B-12 10 μg and folic acid 0, B<sub>0</sub>F<sub>200</sub> = B-12 0 and folic acid 200 μg, B<sub>2</sub>F<sub>200</sub> = B-12 2 μg and folic acid 200 μg, B<sub>10</sub>F<sub>200</sub> = B-12 10 μg and folic acid 200 μg].

**Table 4:** Supplementation Study: Proportion of participants in epidemiological B-12 category of cB12 at baseline, 4, and 12 months.

|  | Baseline | 4 months | 12 months |
| --- | --- | --- | --- |
| **Placebo group** |  |  |  |
| B-12 Adequacy | 46 % | 42 % | 50 % |
| Transitional B-12 status | 52 % | 52 % | 48 % |
| Low B-12 status | 2 % | 6 % | 2 % |
| **B-12, 2µg-group** |  |  |  |
| B-12 Adequacy | 41.5 % | 66 % | 73.3 % |
| Transitional B-12 status | 54.3 % | 34 % | 26.7 % |
| Low B-12 status | 4.2 % | 0 % | 0 % |
| **B-12, 10µg-group** |  |  |  |
| B-12 Adequacy | 35.7 % | 82.7 % | 85.7 % |
| Transitional B-12 status | 60.2 % | 17.3 % | 14.3 % |
| Low B-12 status | 4.1 % | 0 % | 0 % |

Numbers are proportions. cB12 was calculated as per S Fedosov (23). [B-12 Adequacy: cB12 >-0.5; Transitional B-12 status: cB12 > -2.5- < -0.5; Low B-12 status: cB12 <-2.5].

The median compliance with supplements at 12 months was 86%, and it was ≥ 80% in 71% of the participants; only 3% were < 50% compliant. Compliance decreased with the increasing duration of supplementation, which was also evident in the declining circulating B-12 concentrations among poor compliers.

#### Predictors of response to supplementation

We considered plasma B-12 concentrations at 4 and 12 months as the participants’ response to supplementation. We analyzed 4- and 12-month responses in a cross-sectional and also in longitudinal model (**Table 5**). The response was inversely associated with age and weight (both highly correlated; therefore, results are shown only for weight) and directly associated with B-12 dose, duration of supplementation, and compliance. In addition, the response was directly associated with the rise in holoTC measured in the absorption study (‘absorption performance’). At 12 months, the largest effect was of B-12 dose (for 2 µg: β (SE): 0.166 (0.057), *p =* 0.004; for 10 µg: 0.411 (0.056), *p* < 0.0001. The effect size of absorption performance (rise in plasma holoTC during CobaSorb test) was 0.002 (0.001), *p* = 0.012 (Table 5).

**Table 5:**
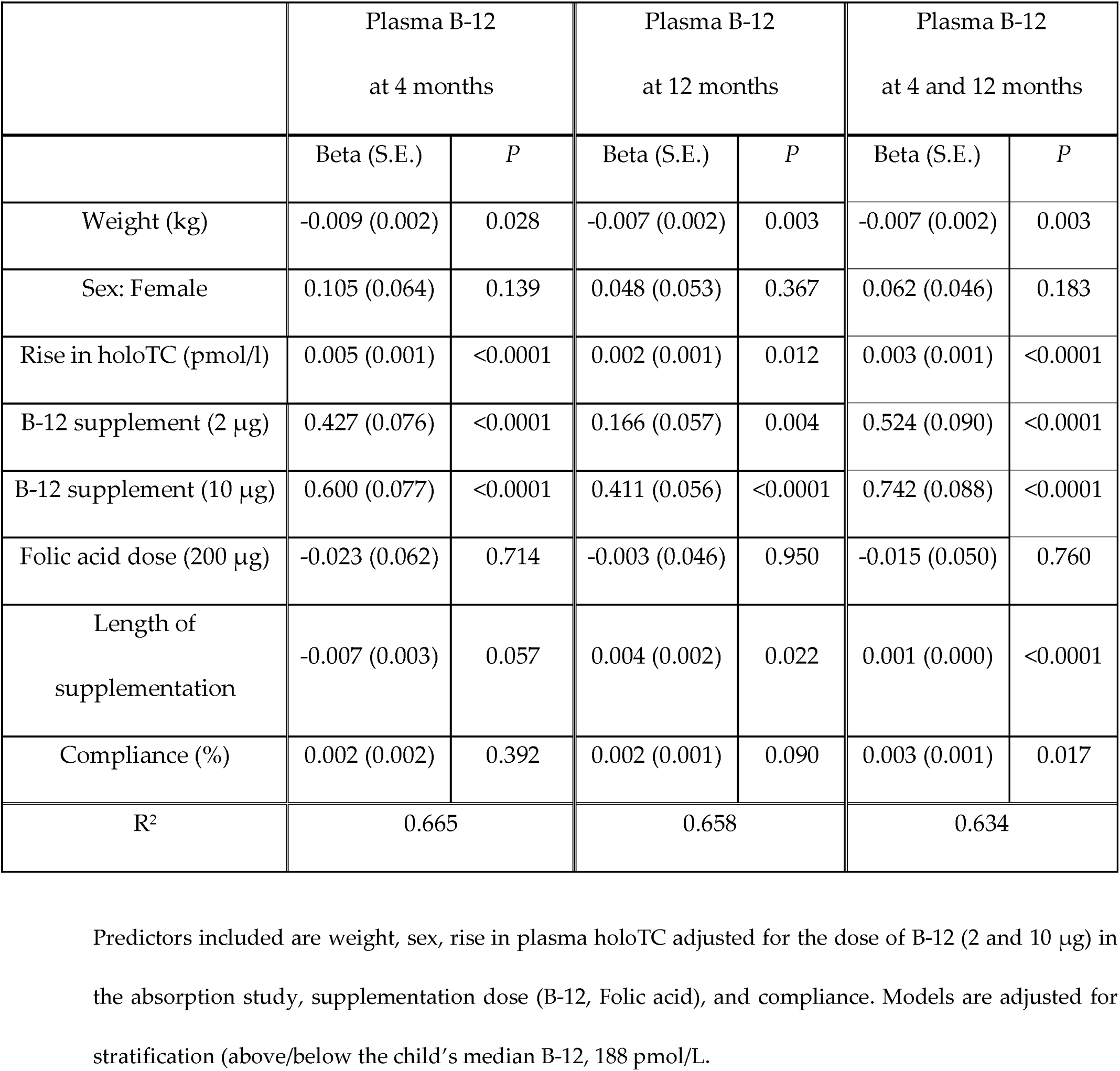
Supplementation Study: Predictors of the response to B-12 supplementation. Outcomes: plasma B-12 at 4 months, 12 months, and 4 + 12 months (longitudinal).

## Discussion

This is a secondary analysis of two linked community-based studies of B-12 metabolism [13, 14] in a longitudinal cohort of largely vegetarian Indian people resident in Pune, India, to expand our understanding of the absorption of B-12 and to study how it relates to response during long-term supplementation.

Our absorption study was the first report from India [13] in a population with a substantial proportion with low plasma B-12 concentrations related to low dietary intake, and the second study was a randomised controlled trial [14] to investigate metabolic effects of physiological doses of B-12 and folic acid in this population. In the CobaSorb absorption protocol, more than 90% absorbed B-12 adequately based on Bor’s original criteria [12] and more than 95% based on the later criteria [24]. Our secondary analysis showed that i) the rise in plasma holoTC concentration (a measure of absorption after an oral challenge with ‘physiological’ dose B-12) is continuously distributed and ii) that it follows the expected physiological rules i.e. the concentration achieved was proportional to the dose and inversely with the weight of the participant. This provides a new understanding because, based on Schilling’s test, there is a tendency among clinicians and researchers to believe in a dichotomous behavior of B-12 absorption (i.e., Yes/No). An additional observation was that the width (spread) of the distribution of rise in holoTC concentration was larger for the 10 µg compared to the 2 µg dose. This suggests that the physiological (carrier-mediated) absorption at near-RDA 2 µg dose is more tightly regulated and therefore less variable, and the extra diffusion-mediated absorption at the higher 10 µg dose is more variable. This understanding could help consider a more ‘personalized’ approach towards dose in clinical practice and public health interventions.

The association of plasma holoTC response with the dose of B-12 and the person’s weight is expected under physiological conditions. A common understanding is that a 5 - 6 µg dose of oral B-12 will saturate the active mechanism. We believe it occurred during the 2 µg x 3 doses experiment. In the 10 µg x 3 dose experiment, it would appear that the active transport mechanisms were saturated, and some passive diffusion took place, which varied between participants and gave the wider distribution in the density plots. It will be interesting for physiologists to model the absorption of B-12 based on such experiments. Recent exciting discoveries using stable isotopes of B-12 are expected to throw further light on this amazingly complex physiological process [6, 25–27].

We extended our secondary analysis to long-term B-12 supplementation to investigate if the performance during the CobaSorb test would predict the response to long-term supplementation. Allowing for body weight, compliance, and dose of B-12, the ‘absorption performance’ was a significant direct predictor. This finding leads us to believe that within physiological limits, the individual’s position within a population ‘tracks’ after an intervention. This again is an important finding for personalized nutrition.

The Schilling test, now obsolete because of restrictions on using radioactive isotopes, had two important requirements: a complete 24-hour urine collection and the presence of normal kidney function [5]. The Schilling test was based on the measurement of radioactivity rather than the concentration of B-12 and was performed mostly in ‘patient’ populations, with fewer controls or volunteers. Assays of circulating B-12 were established by the late 1980s and were used in different absorption protocols. Henze et al [28] described the distribution of serum B-12 concentration after an oral, supraphysiological dose of 1 mg B-12 in 32 normal participants (age range 22 – 75 y) as “Gaussian”. A substantial component of the response would have come from the diffusion of the vitamin across the gastrointestinal mucosa rather than the physiological pathway of absorption. A better understanding of the physiology of B-12 absorption and binding with transport proteins led to the use of holoTC measurement in a B-12 absorption protocol [4, 12, 24]. Bor et al stressed that the dose of B-12 (9 µg x 3 doses) in the CobaSorb test is still near-physiological [12, 24]. The use of an even more physiological dose (2 µg x 3 doses) in a proportion of our subjects further supports the use of a rise in circulating holoTC concentration to study B-12 absorption [13]. The protocol could be modified to be used at home. Importantly, it uses routine laboratory facilities without the requirement of radioactivity (^14^C-labeled B12) or expensive and rare isotopes (^13^C-labeled B12), which depend on a special infrastructure limited to a few laboratories [6, 25–27, 29, 30].

The results of the long-term supplementation using two near physiological doses of B-12 revealed that the performance during the CobaSorb test contributes to the response both at 4 and 12 months. This analysis was possible only because the same cohort participated in both experiments, there were only a few drop-outs, and there was high compliance with the supplementation. We could not find any published studies investigating similar predictors of response to supplementation to compare our results. It was rewarding to see that physiological dose supplementation improved B-12 status satisfactorily within 4 months and persisted at 12 months; the 2 µg dose also performed impressively, which encouraged us to set up a primordial trial [16] in the PMNS cohort.

### Strengths and limitations

Both trials of B-12 (oral absorption and supplementation) were conducted in the same cohort, which belonged to a community where high B-12 insufficiency and poor dietary B-12 intake are prevalent [8, 10]. Studies were population-based, in free-living individuals who were otherwise healthy, and included children as well as adults with a balanced sex ratio. The sample size was large, and the doses of B-12 used were ‘physiological’. The large sample size, the inclusion of children as well as adults, and equal gender distribution add to the strength of the statistical interpretations. On the other hand, this was a very specific population with intergenerational vegetarian practices, which may limit application to the general population.

In summary, secondary analysis of a B-12 absorption study and a 12-month supplementation trial with physiological doses in a community with low circulating B-12 concentrations provided novel findings. The absorption study revealed that the distribution of the absorption characteristics was continuous and not binary and that it followed physiological expectations.

Long-term supplementation study showed that the absorption performance of the participant was a predictor of response to supplementation. Our findings expand the knowledge about B-12 absorption and provide a basis for further studies using modern methods and special populations.

## Data Availability

The raw data supporting the conclusions of this article will be made available upon reasonable request and adequate institutional permissions.

## Acknowledgements

We thank K. J. Coyaji, Director of the KEM Hospital, Pune, for providing the required facilities. We are grateful to the participants of the Pune Maternal Nutrition Study for taking part in these studies. RW is supported by a senior research fellowship from the Council of Scientific and Industrial Research, India CY conceived the idea of this secondary data analysis. UD, ER, and LP wrote the first draft of the manuscript. SB, RW, OD, ER, CY, and UD were involved in the analysis and interpretation of the results. All authors contributed to the article and approved the submitted version.

## Declaration of Generative AI and AI-assisted technologies in the writing process

During the preparation of this work, the authors used the free version of Grammarly to improve language and readability. After using this tool, the authors reviewed and edited the content as needed and take full responsibility for the content of the publication.

## Abbreviations

B-12: Vitamin B_12_
cB12: combined B-12 marker
HoloTC: Holo-transcobalamin
IF: Intrinsic factor
MMA: Methylmalonic acid
PMNS: Pune Maternal Nutrition Study
SAM: S-adenosyl methionine
tHcy: Total homocysteine

